# Monitoring emergence of SARS-CoV-2 B.1.1.7 Variant through the Spanish National SARS-CoV-2 Wastewater Surveillance System (VATar COVID-19) from December 2020 to March 2021

**DOI:** 10.1101/2021.05.27.21257918

**Authors:** Albert Carcereny, Adán Martínez-Velázquez, Albert Bosch, Ana Allende, Pilar Truchado, Jenifer Cascales, Jesús L Romalde, Marta Lois, David Polo, Gloria Sánchez, Alba Pérez-Cataluña, Azahara Díaz-Reolid, Andrés Antón, Josep Gregori, Damir Garcia-Cehic, Josep Quer, Margarita Palau, Cristina González Ruano, Rosa M Pintó, Susana Guix

**Author notes:** Address correspondence to Susana Guix and Rosa Pintó. Equally contributed (Albert Carcereny, Adán Martínez-Velázquez).

## Abstract

**Background:** Since its first identification in the United Kingdom in late 2020, the highly transmissible B.1.1.7 variant of SARS-CoV-2, become dominant in several European countries raising great concern.

**Aim:** The aim of this study was to develop a duplex real-time RT-qPCR assay to detect, discriminate and quantitate SARS-CoV-2 variants containing one of its mutation signatures, the ΔHV69/70 deletion, to trace the community circulation of the B.1.1.7 variant in Spain through the Spanish National SARS-CoV-2 Wastewater Surveillance System (VATar COVID-19).

**Results:** B.1.1.7 variant was first detected in sewage from the Southern city of Málaga (Andalucía) in week 20_52, and multiple introductions during Christmas holidays were inferred in different parts of the country, earlier than clinical epidemiological reporting by the local authorities. Wastewater-based B.1.1.7 tracking showed a good correlation with clinical data and provided information at the local level. Data from WWTPs which reached B.1.1.7 prevalences higher than 90% for ≥ 2 consecutive weeks showed that 8.1±1.8 weeks were required for B.1.1.7 to become dominant.

**Conclusion:** The study highlights the applicability of RT-qPCR-based strategies to track specific mutations of variants of concern (VOCs) as soon as they are identified by clinical sequencing, and its integration into existing wastewater surveillance programs, as a cost-effective approach to complement clinical testing during the COVID-19 pandemic.

## INTRODUCTION

Environmental surveillance of specimens contaminated by human faeces is used to monitor enteric virus disease transmission in the population, and several countries have implemented SARS-CoV-2 wastewater monitoring networks to inform decision making during the COVID-19 pandemic (1–3). In Spain, a nation-wide COVID-19 wastewater surveillance project (VATar COVID-19) was launched in June 2020 (https://www.miteco.gob.es/es/agua/temas/concesiones-y-autorizaciones/vertidos-de-aguas-residuales/alerta-temprana-covid19/default.aspx), and has weekly monitored SARS-CoV-2 levels in untreated wastewater from initially 32 wastewater treatment plants (WWTPs) since then. On March 2021, the European Commission adopted a recommendation on a common approach to establish and make greater use of systematic wastewater surveillance of SARS-CoV-2 as a new source of independent information on the spread of the virus and its variants in the European Union(4). In situations with low or absent SARS-CoV-2 circulation in the community, wastewater surveillance has proven a useful tool as an early warning system (5– 9), and several studies have also tried to infer disease incidence in a community, independent of diagnostic testing availability based on SARS-CoV-2 wastewater concentrations, with considerable uncertainties (10–12).

Despite titanic efforts based on confinement measures and mass-vaccination programs, the emergence of novel variants of concern (VOCs), mainly B.1.1.7, B.1.351 B.1.1.28.1 and recently B1.617.2, so far, suggests that continued surveillance is required to control the COVID-19 pandemic in the long run. Since January 2021, countries within and outside Europe have observed a substantial increase in the number and proportion of SARS-CoV-2 cases of the B.1.1.7 variant, first reported in the United Kingdom (13,14). Since B.1.1.7 variant has been shown to be more transmissible than the previously predominant circulating variants and infections may be more severe (15), countries where the variant has spread and become dominant are concerned on whether the occurrence of the variant will result in increases in total COVID-19 incidence, hospitalizations, and excess mortality due to overstretched health systems.

The emergence of SARS-CoV-2 variants that may increase transmissibility and/or immune escape, points to an imperative need for the implementation of targeted surveillance methods. While sequencing should be the gold standard for variant characterization, cost-effective molecular assays, which could be rapidly established and scaled up may offer several advantages and provide valuable quantitative information without delay.

This study included the development and validation of a one-tube duplex quantitative real-time RT-PCR (RT-qPCR) assay to detect, discriminate and quantitate SARS-CoV-2 variants containing the ΔHV69/70 deletion from variants lacking it, using allelic discrimination probes. Confirmatory sequencing of a subset of samples was performed to be able to ascertain the validity of these assays to trace the community circulation of the B.1.1.7 variant. The RT-qPCR-based assay improved the current variant tracking capability and could be easily implemented for monitoring the emergence of ΔHV69/70 containing SARS-CoV-2 variants (mainly B.1.1.7) in Spain through the nation-wide wastewater surveillance network.

## METHODS

### Wastewater sampling

Influent water grab samples were weekly collected from 32 WWTPs located in 15 different Autonomous Communities in Spain, from middle December 2020 to end of March 2021 (last week of 2020 was not sampled). All samples were transported on ice to one of the 4 participating laboratories of analysis (A, B, C, D), stored at 4°C and processed within 1-2 days upon arrival.

### Sample concentration, nucleic acid extraction and process control

200 ml wastewater samples were concentrated by aluminum hydroxide adsorption-precipitation method, as previously described (7,16), and concentrates were resuspended in 1-2 ml of phosphate buffered saline (PBS). All samples were spiked with a known amount of an animal coronavirus used as a process control virus. Animal coronaviruses differed between participant laboratories and included the attenuated PUR46-MAD strain of Transmissible Gastroenteritis Enteric Virus (TGEV) (17); Porcine Epidemic Diarrhea Virus (PEDV) strain CV777 (kindly provided by Prof. A. Carvajal from University of Leon) and Murine Hepatitis Virus (MHV) strain ATCC VR-764 (18). Nucleic acid extraction from concentrates was performed from 300 µl using the Maxwell® RSC PureFood GMO and Authentication Kit (Promega Corporation, Madison, US), or 150 µl using the NucleoSpin RNA virus kit (Macherey-Nagel GmbH & Co., Düren, Germany), following the manufacturer’s instructions. Each extraction included a negative control and a process virus control used to estimate the virus recovery efficiency. RT-qPCR for process control viruses (19,20) were performed as previously described (7,8). Parallel to ISO 15216-1:2017 (21) for the determination of norovirus and hepatitis A virus in the food chain, samples with a virus recovery ≥ 1% were considered acceptable.

### SARS-CoV-2 RT-qPCR assays

The N1 assay targeting a fragment of the nucleocapsid gene, as published by US CDC (US-CDC 2020), was used to quantify SARS-CoV-2 RNA in the sewage samples, using PrimeScript™ One Step RT-PCR Kit (Takara Bio, USA) and 2019-nCoV RUO qPCR Probe Assay primer/probe mix (IDT, Integrated DNA Technologies, Leuven, Belgium). Different instruments were used by different participating labs, including CFX96 BioRad, LightCycler 480 (Roche Diagnostics, Germany), Stratagene Mx3005P (Applied Biosystems, USA) and QuantStudio 5 (Applied Biosystems, USA).

The S gene was analyzed by a duplex gene allelic discrimination TaqMan RT-qPCR assay, using 400 nM of the following primers targeting the S gene (For-S21708 5’ATTCAACTCAGGACTTGTTCTTACCTT3’ and Rev-S21796 5’TAAATGGTAGGACAGGGTTATCAAAC3’), and 200 nM of the following probes (S_Probe6970in 5’FAM-TCCATGCTATACATGTCTCTGGGACCAATG BHQ1-3’ and S_Probe6970del 5’HEX-TTCCATGCTATCTCTGGGACCAATGGTACT BHQ1-3’). RT-qPCR mastermix was prepared using PrimeScript™ One Step RT-PCR Kit (Takara Bio, USA), and temperature program was 10 min at 50ºC, 3 min at 95ºC, and 45 cycles of 3 seconds at 95ºC and 30 seconds at 60ºC. RT-qPCR analysis for each target included the analysis of duplicate wells containing undiluted RNA, and duplicate wells containing a ten-fold dilution to monitor the presence of inhibitors. Every RT-qPCR assay included 4 wells corresponding to negative controls (2 nuclease-free water and 2 negative extraction controls). Commercially available Twist Synthetic SARS-CoV-2 RNA Controls (Control 2, MN908947.3; and Control 14, EPI_ISL_710528) were used to prepare standard curves for genome quantitation. Both synthetic RNA controls were quantified by droplet-based digital PCR using One-Step RT-ddPCR Advanced kit for probes in a QX200™ System (Bio-Rad), to estimate the exact concentration of genome copies (GC)/µl, prior to construction of RT-qPCR standard curves. Limit of detection (LOD) and limit of quantification (LOQ) were determined for each specific target by running a series of dilutions of the target with 4-10 replicates per dilution. Parameters of all standard curves and estimated LOD and LOQ for the 4 participating laboratories are summarized in supplementary **Table S1**.

### RT-qPCR data analysis and interpretation

The following criteria were used to estimate SARS-CoV-2 gene viral titers. For each specific target, Cq values ≤40 were converted into GC/L using the corresponding standard curve and volumes tested. Occurrence of inhibition was estimated by comparing average viral titers obtained from duplicate wells tested on undiluted RNA with duplicate wells tested on ten-fold diluted RNA. Inhibition was ascertained when difference in average viral titers was higher than 0.5 log_10_, and viral titers inferred from the ten-fold RNA dilution. The percentage of SARS-CoV-2 genomes containing the ΔHV69/70 deletion within the S gene was calculated using the following formula: % = GC/L (Probe6970del) / [GC/L (Probe6970del) + GC/L (Probe6970in)] x 100. In cases with one of the concentrations <LOQ, percentage was calculated using the corresponding LOQ of the assay. Data with both concentrations <LOQ were not considered.

### S gene sequencing and single nucleotide polymorphism (SNPs) identification

Full-length S gene sequencing of wastewater samples was performed following the ARTIC Network protocol (https://artic.network/ncov-2019, with minor modifications) with selected v3 primers (Integrated DNA Technologies) for genome amplification and KAPA HyperPrep Kit (Roche Applied Science) for library preparation (22). Libraries were loaded in MiSeq Reagent Kit 600v3 cartridges and sequenced on MiSeq platform (Illumina). The raw sequenced reads were cleaned from low-quality segments, and mapped against the Wuhan-Hu-1 reference genome sequence to find out variant-specific signature mutations (mutations and indels).

## RESULTS

### Duplex SARS-CoV-2 S gene allelic discrimination RT-qPCR assay validation

To confirm the ability of the RT-qPCR assay to discriminate and estimate the proportion of targets containing the ΔHV69/70 deletion in different scenarios, 9 preparations containing 90:10, 50:50 or 10:90 proportions of B.1.1.7 and Wuhan-Hu-1 synthetic control RNAs at 3 different total concentration levels (1×10^4^, 1×10^3^ and 1×10^2^ GC/rxn) were made and analyzed (**Figure 1**). Assays performed using B.1.1.7 and Wuhan-Hu-1 synthetic control RNAs did not show cross-reactivity (data not shown). Results showed the ability of the method to detect and quantitatively discriminate sequences containing the ΔHV69/70 deletion from wildtype sequences in mixed samples through a wide range of total RNA concentrations often observed in extracted wastewater samples.

**Figure 1.**
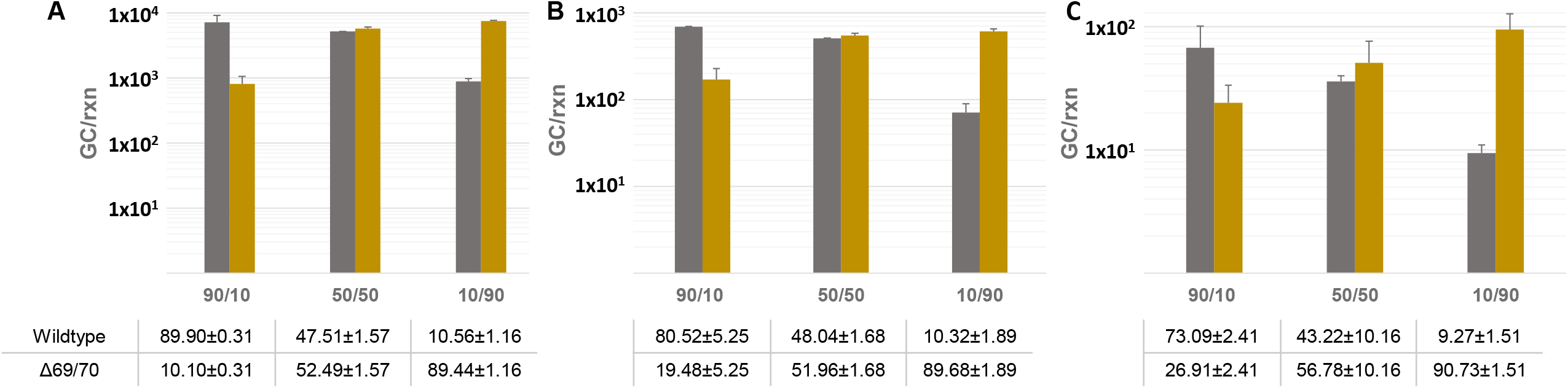
Estimated genome copies (GC) corresponding to wild type SARS-CoV-2 sequences without ΔHV69/70 deletion (grey bars) and sequences containing ΔHV69/70 deletion in the S gene (yellow bars), from 9 preparations at 3 different total concentration levels (**A**: 1×10^4^ GC/rxn, **B**: 1×10^3^ GC/rxn and **C**: 1×10^2^ GC/rxn), and 3 different proportions of Wuhan-Hu-1 and B.1.1.7 GC (90:10, 50:50, and 10:90). Data correspond to mean values ± standard deviations from duplicate samples.

Additionally, to confirm that sequences detected in natural samples collected during the study period containing the ΔHV69/70 deletion corresponded to the B.1.1.7 variant, a subset of 8 samples (4 from week 21_11 and 4 from week 21_12), with B.1.1.7 proportion estimates ranging between 35% and 100% were analyzed by NGS sequencing of the S gene. Variant analysis showed a total of 12 nucleotide substitutions and 4 deletions in comparison with the reference genome of SARS-CoV-2 isolate Wuhan-Hu-1 (MN908947.3), most of them being specific for B.1.1.7 variant. No strong correlation was observed between coverage and specific amplicons. Between 3 and 7 mutation markers out of 9 markers specific for B.1.1.7 variants in the spike region (ΔHV69-70, ΔY144, N501Y, A570D, D614G, P681H, T716I, S982A and D1118H) were detected in all samples, confirming that the RT-qPCR assay could be used to trace the occurrence of B.1.1.7 variant, as suggested by the recently published EU recommendation (4) (**Figure 2**). Seven additional amino acid substitutions/deletions, which were not specific for B.1.1.7 variant, were also detected: G142V, A222V, G257V, S375P, ΔT376, F377L and K537E. Of these substitutions, as of April 28^th^, G142V and G257V had already been reported in 1128 and 259 sequences published at GISAID database, respectively, but the others had only been reported at low frequencies. Substitutions S375P/ΔT376/F377L affecting 3 consecutive residues within the Receptor Binding Domain (RBD) and identified in 35% of sequences from WWTP-31 in Madrid, have been individually reported less than 15 times in several countries, including Spain. However, our data report the occurrence of the 3 mutations within the same variant. Of note, mutations in these residues have been related to antigenic drift (23,24). Substitution K537E had been reported once in a nasopharyngeal specimen belonging to B.1.177 (EPI_ISL_1547898; hCoV-19/Slovakia/UKBA-2586/2021). Marker A222V is also present in B.1.777 variant, which originated in Spain became the predominant variant in most European countries during the second pandemic wave (25). Sequence data obtained in this study are available at Genbank (SAMN19107574 and PRJNA728923).

**Figure 2.**
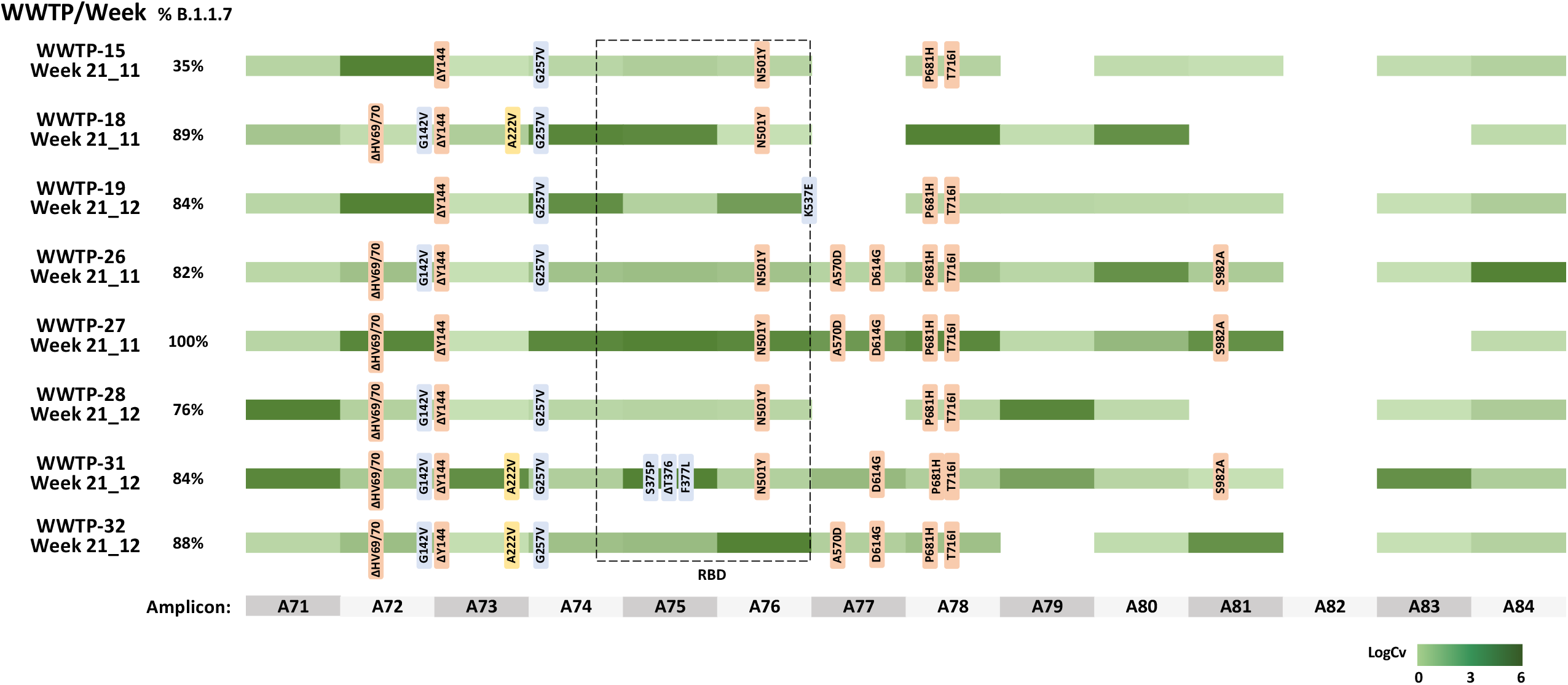
Overview of the nucleotide substitutions detected in SARS-CoV-2 S gene sequences from wastewater samples (n=8) as compared to the SARS-CoV-2 isolate Wuhan-Hu-1 reference genome (MN908947.3). B.1.1.7-specific markers are shown in light orange; yellow markers show mutations described in B.1.777 variant, and blue markers indicate others. RBD (Receptor Binding Domain) is indicated with a dotted square. Amplicon numbers are shown at the bottom. Shaded green colors indicate sequence coverage in logarithmic scale for each amplicon.

### Temporal and geographical emergence of B.1.1.7 variant in the Spanish territory

Wastewater samples from 32 Spanish WWTPs from mid December 2020 to end of March 2021 were weekly analyzed to monitor the emergence of B.1.1.7 in the territory. Total levels of SARS-CoV-2 RNA were determined by RT-qPCR using N1 target as well as the S discriminatory RT-qPCR, without normalization by the population number (**Figure 3**). A moderate correlation was observed between both SARS-CoV-2 genome concentration measures between N1 and total S gene titers (R^2^=0.303, data not shown).

**Figure 3.**
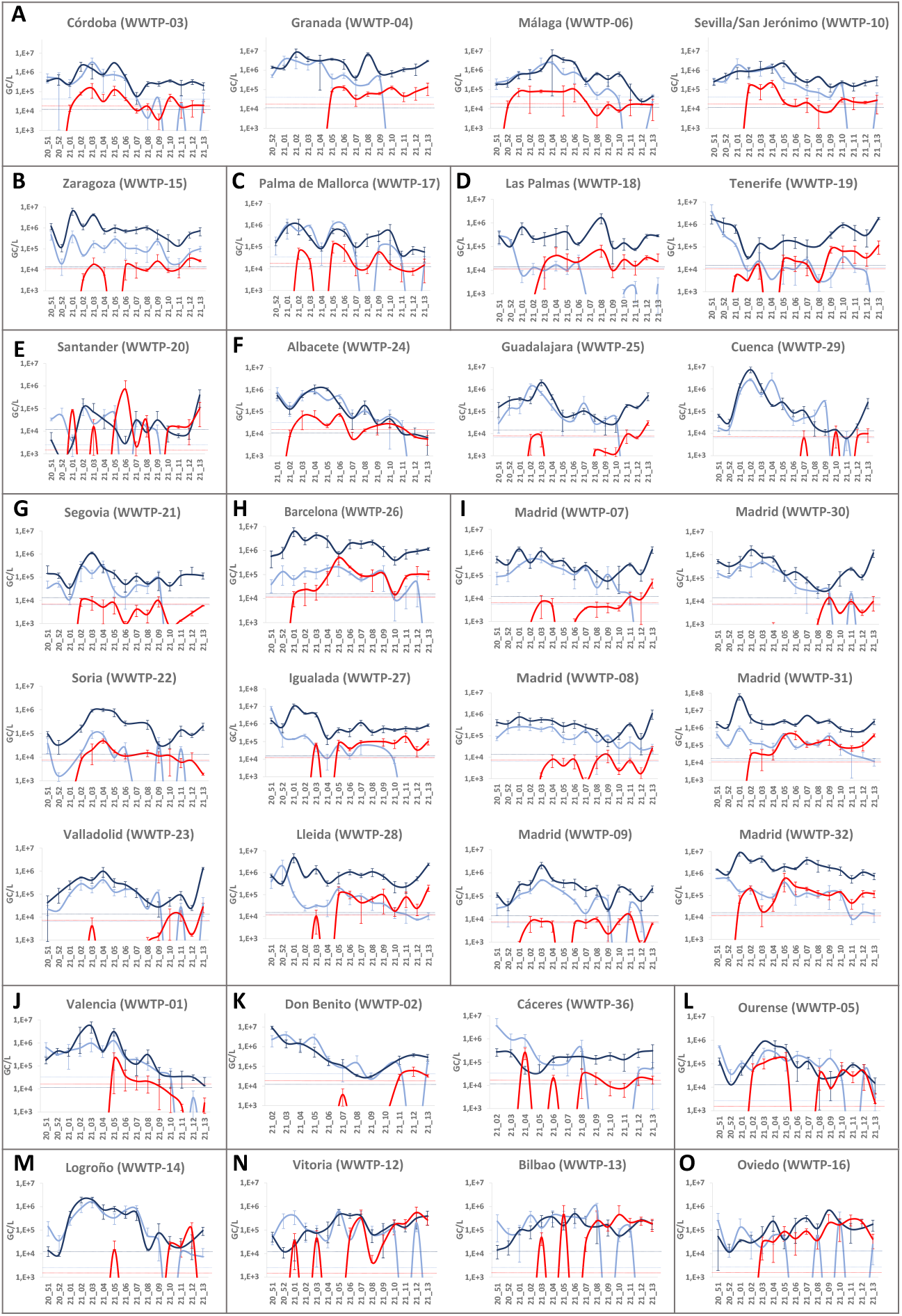
Concentration of SARS-CoV-2 RNA in wastewater samples collected in Spain from December 2020 to March 2021, as measured by N1 RT-qPCR (dark blue), and duplex S gene allelic discrimination RT-qPCR [wildtype S (light blue) and B.1.1.7 S (red)]. Wastewater treatment plants (WWTPs) are alphabetically grouped by Autonomous Communities in Spain (**A**: Andalucía, **B**: Aragón, **C**: Baleares, **D**: Canarias, **E**: Cantabria, **F**: Castilla-La Mancha, **G**: Castilla y León, **H**: Cataluña, **I**: Com. De Madrid, **J**: Com. Valenciana, **K**: Extremadura, **L**: Galicia, **M**: La Rioja, **N**: País Vasco, **O**: Pr. Asturias). Data represent average values and error bars standard deviation. Dotted lines correspond to the limit of quantification of assays.

Lockdown measures in Spain during the study period were remarkable (mandatory use of face mask, nighttime curfews, restrictions regarding bar and restaurant opening times, social gathering restrictions, restricted opening hours and attendance, municipality of residence confinement implemented in most regions, etc.) and were associated to the nationwide State of Alarm, in place since October 2020. As most European countries, at the clinical level, a peak in COVID-19 cases occurred between the end of December 2020 (week 20_52) and early February 2021 (week 21_05). As measured through the N1 target, a peak in SARS-CoV-2 genome levels in wastewater was observed in week 21_01 in 9 regions including Zaragoza (**Fig 3B**; WWTP-15), Las Palmas in Canary Islands (**Fig 3D**; WWTP-18), 3 cities in Catalonia (**Fig 3H**; WWTP-26, 27 and 28) and Madrid (**Fig 3I**; WWTP 07, 08, 31 and 32); in week 21_02 in regions including Córdoba and Granada in Andalucía (**Fig 3A**; WWTP03 and WWTP-04), Palma de Mallorca (**Fig 3C**; WWTP-17), Santander (**Fig 3E**; WWTP-20), Cuenca (**Fig 3F**; WWTP-29), Madrid (**Fig 3I**; WWTP-30) and Logroño (**Fig 3M**; WWTP-14); in week 21_03 in Guadalajara (**Fig 3F**; WWTP-25), Segovia (**Fig 3G**; WWTP-21), Soria (**Fig 3G**; WWTP-22), Madrid (**Fig 3I**; WWTP-09), Valencia (**Fig 3J**; WWTP-01) and Ourense (**Fig 3L**; WWTP-05); in week 21_04 in Málaga (**Fig 3A**; WWTP_06), Albacete (**Fig 3F**; WWTP-24), Valladolid (**Fig 3G**; WWTP-23), Bilbao (**Fig 3N**; WWTP-13) and Oviedo (**Fig 3O**; WWTP-16) and, and in week 21_05 in Sevilla (**Fig 3A**; WWTP-10) and Vitoria (**Fig 3N**; WWTP-12). SARS-CoV-2 genome titers in Tenerife (Canary Islands) (**Fig 3D**; WWTP-19), were already at peak titers in the first week of the study (**Figure 3**).

First detection of B.1.1.7 variant in wastewater samples occurred in the Southern city of Málaga (Andalucía) in week 20_52 (**Figure 4**). The first week of January 2021, it could also be detected in the 2 largest cities (Madrid and Barcelona), in 2 Northern cities (Santander and Vitoria), another city in Andalucía (Córdoba) and in Tenerife (Canary Islands), suggesting that multiple introductions occurred during Christmas holidays in different parts of the Spanish territory, and representing 20% of all sampled WWPTs. B.1.1.7 levels detected in week 21_01 were lower than 10% in most WWTPs, with the exception of WWTP-32 in Madrid, which was of 22.9%. Percentages of WWTPs with B.1.1.7 detection increased progressively, up to 56% by week 21_04, 91% by week 21_08, and 97% by week 21_13. Our data also showed that while total level of SARS-CoV-2 RNA genomes measured by N1 target showed a slight decline for several weeks after the first peak observed during early 2021, this negative trend was slowed or reversed in most WWTPs from the time when the proportion of B.1.1.7 became more abundant (**Figure 3**). In 8 cities, a significant increase of N1 RNA titers higher than 0.7 log_10_ with respect to preceding week was observed in the last week of the study (**Fig 3D** WWTP-19, **Fig 3E** WWTP-20, **Fig 3F** WWTP 29, **Fig 3G** WWTP 23, **Fig3H** WWTP 28, **Fig 3I** WWTP07, 08 and 30).

**Figure 4.**
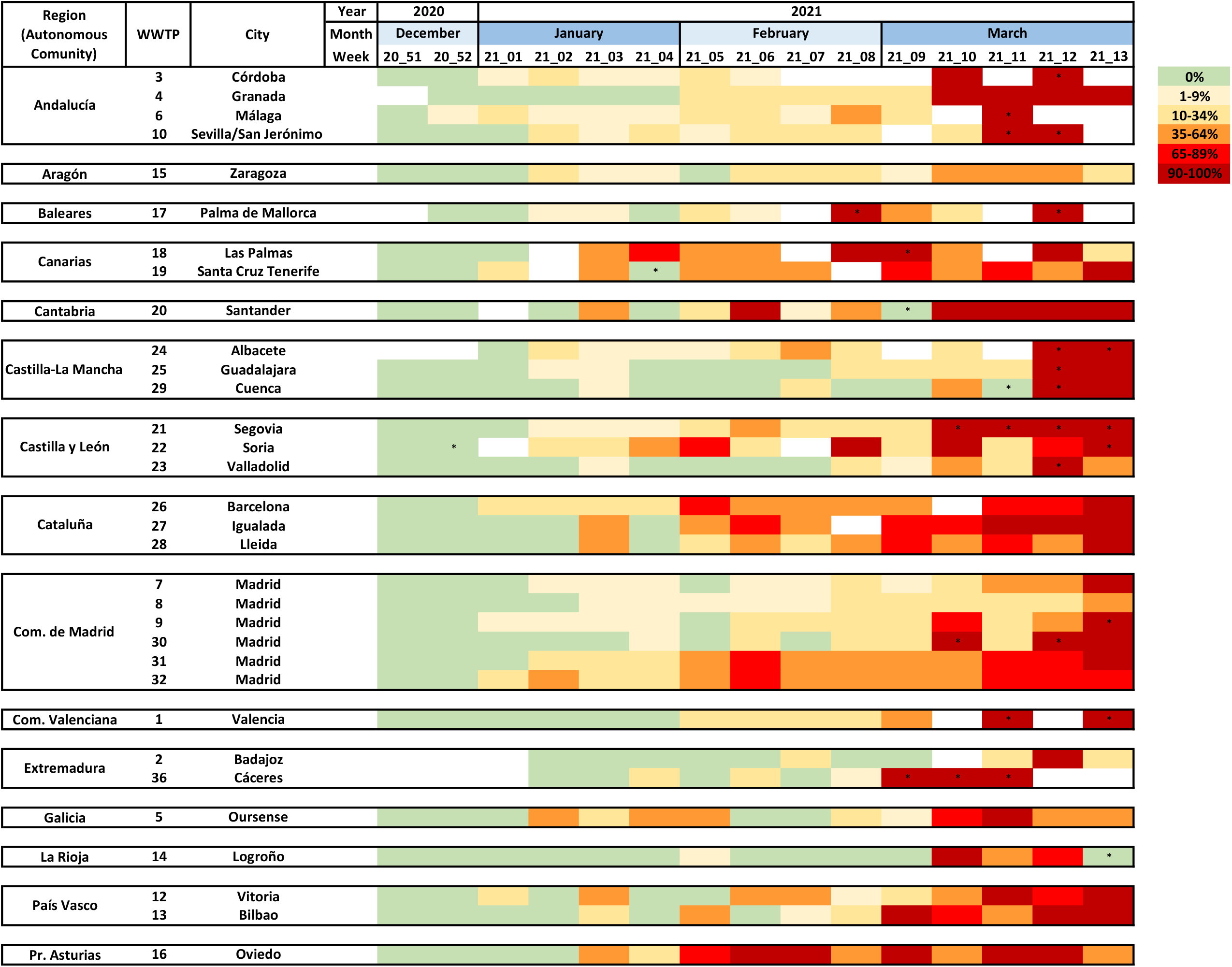
Evolution of B.1.1.7 SARS-CoV-2 prevalence over time, as measured by duplex RT-qPCR in wastewater samples from 32 wastewater treatment plants (WWTPs). As in Figure 3, data are alphabetically shown according to Autonomous Community. * indicates samples with detection of a single variant, but with titers <LOQ.

The relative proportion of the B.1.1.7 variant in wastewater could be estimated for 91% of positive samples. **Figure 4** shows the heatmap of the evolution of B.1.1.7 prevalence in wastewater over time. Predominance of B.1.1.7 variant with prevalences ≥90% was reached in all WWTPs except in WWTP 9 and 32 in Madrid, and WWTP-15 (Zaragoza). In Madrid B.1.1.7 reached 48% and 86%, and in Zaragoza 51% at the end of the study period, although it kept progressively increasing thereafter (data not shown). When considering data from 11 WWTPs, which showed at least 2 consecutive weeks with B.1.1.7 percentages near fixation (90-100%) as a confirmation of predominance (**Table S2**), we could estimate that approximately 8.1±1.8 weeks-time were required for B.1.1.7 variant to become predominant in sewage. This would correspond to an average increase rate of 11.7% (9.6-15.1%) per week.

The proportion of B.1.1.7 was compared to the prevalence detected at the clinical level. The abundance of B.1.1.7, as a fraction of all sequenced clinical specimens by the local authorities in each Autonomous Community, was obtained from update reports of the epidemiological situation of the variants of SARS-CoV-2 of importance published by the Spanish Ministry of Health (26). When comparing the proportion of B.1.1.7 estimated from wastewater with the proportion estimated from sequencing of clinical isolates, a good correlation was observed (R^2^=0.5012) (**Figure 5A**). On a geographical/temporal analysis, wastewater testing allowed to confirm circulation of B.1.1.7 variant before its identification in clinical specimens. Data for 3 selected weeks are shown in **Figure 5B**. By week 21_04 at the end of January, at the clinical level the variant was only detected, on clinical samples, in Galicia and País Vasco, mainly due to the low number of sequenced isolates in most regions, while it was detected in 18/32 (56%) WWTPs. At the end of the study, all Autonomous Communities Public Health departments reported percentages higher than 60%, but while some WWTPs showed very high percentages some others showed relatively lower proportions, indicating differences at the local level.

**Figure 5.**
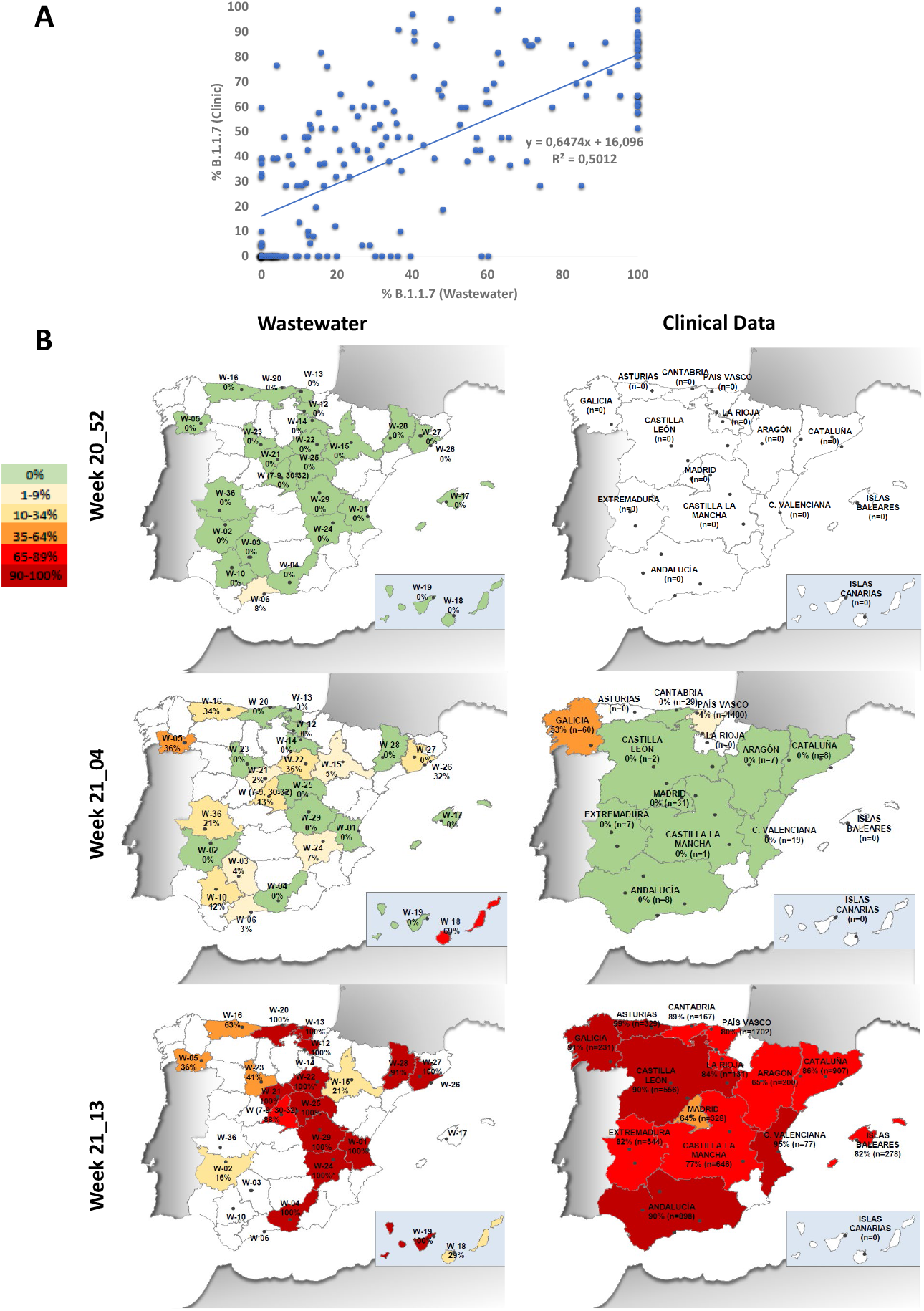
Comparison of B.1.1.7 estimates from wastewater testing and clinical epidemiological surveillance. **(A)** Correlation between B.1.1.7 proportions estimated by duplex RT-qPCR from wastewater and data reported by local authorities from clinical specimens sequencing. **(B)** Geographic and temporal evolution of B.1.1.7 SARS-CoV-2 emergence in Spain during the study period, estimated from wastewater samples (left panels) and reported in clinical data (right panels). For wastewater data, percentages are indicated for each WWTP. * indicates samples with detection of a single variant, but with titers <LOQ. For clinical data, percentages are indicated for each Autonomous Community and number in parenthesis indicates the number of cases under sequence study during that week. Communities in for which data were not available are depicted colorless.

## DISCUSSION

As a cost-effective approach to screen thousands of inhabitants, wastewater-based epidemiology (WBE) is a valuable tool to anticipate the circulation of specific pathogens in a community and to closely track their incidence evolution through space and time (27,28). This surveillance has proven to be extremely useful to monitor the circulation of total SARS-CoV-2 in different parts of the world (1–3), and may be effective for tracking novel SARS-CoV-2 VOC. Despite NGS would allow the definitive identification of specific variants and has already been applied on sewage samples (29,30), it is resource and time-consuming, thus limiting the number of samples that can be processed and the number of labs which can implement this approach on a regular basis. In addition, most deep sequencing protocols allow the identification of signature mutations within short individual reads, which when detected in wastewater samples containing a mixture of different isolates, may not be proof of co-occurrence of such mutations within the same genome, thus providing only an indirect evidence of the presence of a certain variant (31). Finally, the use of NGS is also challenged by the presence of inhibitors in samples, limiting its success rate and depth coverage. Given its higher tolerance to inhibitors, droplet digital RT-PCR has been acknowledged as a suitable approach to simultaneous enumerate the concentration of variants with the N501Y mutation and wildtype in wastewater (32), but droplet digital RT-PCR widespread use may be limited nowadays by the high economic investment in instrumentation. On the other hand, the use of RT-qPCR methods offers the advantage of rapid turnaround time, lower cost, and immediate availability in most public health laboratories. In the current study, we validated a duplex RT-qPCR assay to discriminate and enumerate SARS-CoV-2 variants containing the ΔHV69/70 deletion from variants lacking it. Among molecular markers specific for B.1.1.7 variant, the 6-nucleotide deletion corresponding to residues 69/70 was chosen because it offers the possibility to design highly-specific robust probes to be used in wastewater samples, which unlike clinical specimens, will contain mixed sequences in most cases. Similar to the TaqPath COVID-19 assay (Thermo Fisher Scientific) and other RT-qPCR protocols designed for clinical diagnosis (33), the novel duplex RT-qPCR assay developed in this study proved highly specific and discriminatory.

The ΔHV69/70 deletion is located within the N-terminal domain of the S glycoprotein and has been described to be located at a recurrent deletion region (RDR), and phylogenetic studies showed that it has arisen independently at least 13 times (34). In addition to being a signature mutation of highly transmissible B.1.1.7 variant, it has also been described in other lineages, including cluster-5 variant, identified both in minks and humans in Denmark, some isolates belonging to 20A/S:439K variant, which emerged twice independently in Europe, B.1.258 and B.1.525 lineages (34–38), although none of these other lineages have been shown to spread widely. According to GISAID, from a dataset of 442,175 sequences collected from 1 December 2020 to 31^st^ March 2021 containing Δ69, as a hallmark of ΔHV69/70 deletion, the proportion of sequences which were classified as B.1.1.7 was of 92.4% (for clinical sequences isolated in Spain during the same period, this percentage was of 98.1%). Among sequences containing Δ69 not classified as B.1.1.7 variant, other lineages including B.1.258, B.1.525, B.1.177, B.1.429+B.1.427, P1, B.1.351 and B.1.617 were observed in a minority of cases. Among sequences belonging to the predominant lineage in Spain at the onset of this study, B.1.177, only 0.23% of sequences deposited in GISAID contained Δ69, confirming that detection ΔHV69/70 is highly indicative of a genome belonging to B.1.1.7 lineage. Finally, in the NGS analysis performed on 8 selected samples with a high proportion of ΔHV69/70 containing genomes, between 3-8 additional B.1.1.7 mutation signatures were identified, confirming that the detected genomes very likely correspond to the B.1.1.7 variant.

During the study period, weekly wastewater estimates of the proportion of B.1.1.7, representing a larger and more comprehensive proportion of typed cases including both symptomatic and asymptomatic cases, well reflected the trends in the reported sequenced clinical cases in most regions. Despite the number of clinical specimens sequenced from public health laboratories was not high during the study period, and showed strong geographic differences, a correlation was observed between the proportion of B.1.1.7 cases observed at the clinical level and data estimated from sewage (**Figure 5A**). Sewage surveillance allowed the identification of B.1.1.7 circulation in the Spanish territory in the Southern city of Málaga before it was confirmed at the clinical level by National Public Health Authorities, and allowed us to infer multiple simultaneous introductions during Christmas and New Year’s holidays in distant parts of the country (Madrid, Barcelona, Santander, Vitoria, Córdoba and Tenerife). By the end of January 2021, only 13% (2/15) Autonomous Communities had reported B.1.1.7 clinical cases, while circulation in sewage had been confirmed in 67% (10/15) of them, confirming its use as an early warning approach. Data from 11 WWTPs which reached B.1.1.7 near fixation rates higher than 90% for ≥ 2 consecutive weeks showed that 8.1±1.8 weeks were required to reach B.1.1.7 predominance, which would be a slightly shorter time than what has been locally observed at the clinical level. A research publication reported first detection of imported B.1.1.7 clinical cases in Madrid in week 20_52 (December 2020), and a proportion of 62% of total newly diagnosed COVID-19 cases 10 weeks later (39). Data from other studies are also the UK reported by ECDC show that B.1.1.7 cases went from less than 5% of all positive cases to more than 60% in less than six weeks during November to mid-December 2020 (40), and Davies et al. demonstrated that it became dominant throughout the country (15). Estimates from the US indicate that B.1.1.7 would become dominant in most states 4 months after its first identification in late November 2020 (14).

Our data also showed that predominance of B.1.1.7 variant correlated with a slowdown in the negative trend of total SARS-CoV-2 wastewater levels, which had been observed from early 2021 in most cities, probably due to lockdown measures and the mass-vaccination campaign, which was initiated the last week of 2020. Even in some cities, including Tenerife (**Fig 3D**; WWTP-19), Santander (**Fig 3E**; WWTP-20), Cuenca (**Fig 3F**; WWTP-29), Valladolid (**Fig 3G**; WWTP-23), Lleida (**Fig 3H**; WWTP-27) and Madrid (**Fig 3I**; WWTP-07, 08 and 30), total SARS-CoV-2 levels showed a positive trend at the end of the study, suggesting that the emergence of B.1.1.7 cases produced a higher transmission rate and a slight increase in COVID-19 incidence, as confirmed by clinical epidemiological data reported by the Spanish Ministry of Health, reporting an incidence peak between the end of March and April 2020 (41). Despite this positive trend markedly observed in some regions, the smooth running of the mass vaccination campaign starting on December 27^th^ 2020, in addition to non-pharmaceutical interventions, likely contributed to minimizing the impact of B.1.1.7 emergence. As of end of March 2021, the percentage of the Spanish population who had been partially immunized or totally vaccinated were of 13.2 % and 6.8 %, respectively.

Finally, despite the state of alarm decreed by the Spanish government, as a measure to unify confinement and restriction measures across the country, was maintained throughout the study period, predominance of B.1.1.7 variant was not homogeneous, and dynamics were variable among cities across the country. For instance, a rapid predominance of B.1.1.7 variant was observed in Granada (WWTP-4) and Cáceres (WWTP-36), while in other cities, B.1.1.7 reached prevalences higher than 90% by week 3-6 after first positive detection and decreased thereafter for 2-3 weeks. Several reasons could explain these waves, including differences in regional social distancing behaviors, repetitive B.1.1.7 case imports, introduction of additional variants, climatic effect on sewage composition, size of WWTP, or variability related to the use of grab samples instead of composite samples.

This study highlights the use of WBE as a cost-effective, non-invasive and unbiased approach which may complement clinical testing during the COVID-19 pandemic, and demonstrates the applicability of duplex RT-qPCR assays on sewage surveillance as a rapid, attractive, and resourceful method to track the early circulation and emergence of known VOC in a population. The current strategy could be readily adaptable to track specific mutations of other VOC as soon as they are identified by clinical genomic sequencing in the future and integrated into existing wastewater surveillance programs.

## Data Availability

The data that support the findings of this study are available from the corresponding authors, upon request.

## ACKNOWLEDGEMENTS

This work was partially supported by the COVID-19 wastewater surveillance project (VATar COVID19), funded by the Spanish Ministry for the Ecological Transition and the Demographic Challenge of and the Spanish Ministry of Health; grants from CSIC (202070E101) and MICINN co-founded by AEI FEDER, UE (AGL2017-82909); grant ED431C 2018/18 from the Consellería de Educación, Universidade e Formación Profesional, Xunta de Galicia (Spain); Direcció General de Recerca i Innovació en Salut (DGRIS) Catalan Health Ministry Generalitat de Catalunya through Vall d’Hebron Research Institute (VHIR), and Centro para el Desarrollo Tecnológico Industrial (CDTI) from the Spanish Ministry of Economy and Business, grant number IDI-20200297. Pilar Truchado is holding a Ramón y Cajal contract from the Ministerio de Ciencia e Innovación. Adan Martinez is holding a predoctoral fellowship FI_SDUR from Generalitat de Catalunya. We gratefully acknowledge all the staff involved in the VATar COVID-19 project, working with sample collection and logistics. The authors are grateful to Promega Corporation (Madison, US) for technical advice, and thank Andrea Lopez de Mota for her technical support.

## AUTHORS’ CONTRIBUTIONS

Study design: Rosa M Pintó, Albert Bosch, Susana Guix, Ana Allende, Gloria Sanchez, Jesus L Romalde. Data generation and interpretation: Albert Carcereny, Adan Martínez-Velázquez, Pilar Truchado, Jenifer Cascales, Marta Lois, David Polo, Alba Pérez-Cataluña, Azahara Díaz-Reolid, Josep Gregori, Damir Garcia-Cehic. Resources: Margarita Palau, Cristina González Ruano. Funding acquisition: Albert Bosch, Ana Allende, Gloria Sánchez, Jesus L Romalde, Andres Anton, Josep Quer. Conceptualization: Rosa M Pintó, Albert Bosch, Susana Guix. Manuscript writing: Rosa M Pintó, Susana Guix. Critically revision of the manuscript: All authors.

