## Supplemental Table1 Table 2 for "Monitoring emergence of SARS-CoV-2 B.1.1.7 Variant through the Spanish National SARS-CoV-2 Wastewater Surveillance System (VATar COVID-19) from December 2020 to March 2021"

SUPPLEMENTARY MATERIAL

This supplementary material is hosted by Eurosurveillance as supporting information alongside the article “**Monitoring emergence of SARS-CoV-2 B.1.1.7 Variant through the Spanish National SARS-CoV-2 Wastewater Surveillance System (VATar COVID-19) from December 2020 to March 2021**”, on behalf of the authors, who remain responsible for the accuracy and appropriateness of the content. The same standards for ethics, copyright, attributions and permissions as for the article apply. Supplements are not edited by Eurosurveillance and the journal is not responsible for the maintenance of any links or email addresses provided therein.

**Table S1**. Parameters defining standard curves and limit of detection (LOD) and limit of quantification (LOQ) (genome copies/reaction, GC/rxn) for N1 target and S duplex RT-qPCR assay, of all 4 participating laboratories.

| **Lab** | **Target** | **Slope** | **Intercept** | **Efficiency** | **R^2^** | **LOD (GC/rxn)** | **LOQ (GC/rxn)** |
| --- | --- | --- | --- | --- | --- | --- | --- |
| A | N1 | -3.696 | 42.45 | 86.45 | 0.981 | 11.23 | 28.45 |
|  | S_Probe6970in | -3.702 | 40.22 | 86.24 | 0.977 | 6.07 | 27.61 |
|  | S_Probe6970del | -3.615 | 40.31 | 89.07 | 0.986 | 6.39 | 24.76 |
| B | N1 | -3.473 | 39.46 | 94.10 | 0.946 | 10.33 | 36.13 |
|  | S_Probe6970in | -3.734 | 41.47 | 85.28 | 0.967 | 8.00 | 21.59 |
|  | S_Probe6970del | -3.430 | 41.29 | 95.69 | 0.989 | 5.60 | 7.92 |
| C | N1 | -3.667 | 39.99 | 87.37 | 0.967 | 8.14 | 12.69 |
|  | S_Probe6970in | -3.867 | 42.47 | 81.38 | 0.996 | 29.58 | 30.60 |
|  | S_Probe6970del | -3.612 | 39.43 | 89.18 | 0.989 | 2.63 | 14.76 |
| D | N1 | -3.169 | 37.38 | 105.85 | 0.977 | 5.74 | 19.90 |
|  | S_Probe6970in | -3.054 | 37.97 | 112.52 | 0.983 | 3.91 | 7.40 |
|  | S_Probe6970del | -3.409 | 38.62 | 96.40 | 0.996 | 5.60 | 15.31 |

**Table S2.** Time (weeks) required for B.1.1.7 variant to reach a 90-100% prevalence for at least 2 consecutive weeks.

| **WWTP** | **City** | **Weeks** |
| --- | --- | --- |
| 4 | Granada | 5 |
| 10 | Sevilla | 9 |
| 13 | Bilbao | 9 |
| 20 | Santander | 7 |
| 21 | Segovia | 8 |
| 24 | Albacete | 10 |
| 25 | Guadalajara | 10 |
| 27 | Igualada | 8 |
| 29 | Cuenca | 10 |
| 30 | Madrid | 8 |
| 36 | Cáceres | 5 |
